# Metformin, monoacylglycerol lipase expression, cognition and emotion recognition in people with multiple sclerosis and comorbid type II diabetes: A case-control study

**DOI:** 10.1101/2024.12.06.24318151

**Authors:** Lisa A. S. Walker, Sanghamithra Ramani, Jordan D. Pumphrey, Tamanna Islam, Jason A. Berard, Matthew Seegobin, Mai Buckle, Jennifer M. Lymer, Mark S. Freedman, Jing Wang

**Author notes:** indicates co-first authors (Lisa A. S. Walker, Sanghamithra Ramani). Corresponding Author: Dr. Lisa Walker The Ottawa Hospital – General Campus 501 Smyth Road Suite 7300 Ottawa, Ontario, Canada K1H 8L6.

## Abstract

**Background:** Diabetes (DM), a common comorbidity, results in poorer cognition in people with multiple sclerosis (PwMS). Metformin may be a treatment option given cognitive benefits. Metformin represses monoacylglycerol lipase (Mgll), accompanied by improvements in cognition in animals.

**Aims:** To determine 1) whether metformin represses Mgll in humans, 2) if Mgll correlates with cognition/emotion recognition, and 3) if cognition differs between groups.

**Methods:** A convenience sample of seventeen PwMS and DM on metformin, 4 with MS and DM not on metformin, 10 with MS, and 21 healthy controls completed BICAMS and measures of premorbid ability, emotion recognition, mood and fatigue. Blood draw established Mgll levels. T-tests determined group differences in Mgll. Correlational analyses examined if Mgll correlated with cognition. ANCOVA evaluated differences in cognition/emotion recognition.

**Results:** Given small samples, we combined groups to determine if metformin impacted Mgll regardless of diabetes status. Significant differences in Mgll (*t* = -2.07, *p* = .05), suggested that metformin suppresses Mgll. No relationship was found between Mgll and cognition/emotion recognition. Differences were found between PwMS and DM compared to controls in verbal learning (*F* = 5.85, *p* = .02) and memory (*F* = 5.62, *p* = .02).

**Conclusions:** Metformin suppresses Mgll in humans suggesting metformin be evaluated as a potential MS treatment. Mgll did not correlate with cognition possibly due to sample size or methodology. Combined impact of MS and DM negatively impacts cognition, supporting literature demonstrating that vascular comorbidity increases risk of cognitive dysfunction. Findings support pursuing clinical trials evaluating metformin efficacy.

## 1. Introduction

Multiple sclerosis (MS) is a progressive neuroinflammatory and demyelinating disease, often leading to neurological disabilities, including impaired mobility, sensation, coordination, cognition, mood and social cognition(1–3). MS is often accompanied by a variety of comorbidities, including cardiovascular risk factors such as type II diabetes mellitus (DM)(4). The prevalence of DM in MS has been increasing over time at a higher rate than in the general population(5). DM accounts for 12.2% of the comorbidities diagnosed during the year following the first diagnosis of MS(6). Outcomes in those with MS and comorbid DM are often worse than those with MS alone(7) and include a more rapid progression in ambulation dysfunction(8) and EDSS progression(9), increased hospitalizations(10), reduced cortical and deep gray matter volumes(11), and increased mortality(12).

An outcome of particular importance to the quality of life of people with MS (PwMS) is cognition. Cognitive impairment affects approximately half of PwMS overall, with prevalence rates increasing to between 80 and 90% in progressive subtypes(13). Emerging literature has demonstrated that comorbid diabetes is a risk factor for worse cognitive outcomes. Diabetes has been associated with lower scores on a screening measure of cognition(14), as well as lower visual memory(15), verbal learning and memory(16), and verbal fluency performance(15, 16). Findings regarding processing speed have been variable with two studies noting poorer performance in those with comorbid diabetes on the written(14) and oral(16) versions of the Symbol Digit Modalities Test, but another study showing poorer performance on only the fourth quartile of the Processing Speed Test(17). Notably, the difference in cognition between those with MS and those with MS and vascular comorbidities disappears when differences in brain structure are accounted for(16). These structural differences have been attributed to several different mechanisms, including molecular alterations in neurons due to prolonged hyperglycemia and hyperinsulinemia(14), increased peripheral inflammation, endothelial injury, and alterations in blood vessel function, cerebral blood flow and metabolism(16).

Given the negative impact of DM on outcomes in PwMS, there is a need for treatments to minimize these effects. Metformin, a first-line medication commonly used to treat DM, was recently identified as a candidate drug for repurposing and testing in clinical trials of people with progressive MS given the anti-inflammatory effects, as well as the potential for promoting remyelination and neuroprotection(18). A recent trial demonstrated that treatment of individuals with MS and metabolic syndrome with metformin resulted in anti-inflammatory effects. These metabolic changes were associated with reduced disease activity on MRI(19). While the impact of metformin on cognitive functioning has been addressed in other populations such as aging adults(20), individuals with Alzheimer’s(21, 22) and those with Huntington’s(23) with varying effects, no published research to date has evaluated the potential impact of metformin on cognition in individuals with MS. Although there are a few large clinical trials of metformin currently ongoing(24, 25), only one has a cognitive outcome(26). Results from animal studies (see below) are promising with regard to cognitive benefits(27), thus, the potential cognitive benefits of metformin in PwMS demand further investigation.

MS is increasingly being considered a metabolic disease(28, 29) in addition to an autoimmune disease. In both PwMS and animal models, the homeostasis of lipid metabolism collapses during the acute-phase inflammatory response. This starts a feedback loop of increased oxidative stress and elevated inflammatory responses(30). Monoacylglycerol lipase (Mgll), which hydrolyses the endocannabinoid 2-arachidonoylglycerol (2-AG) into arachidonic acid (ARA), has attracted much attention as a therapeutic target for MS treatment(31–33). The altered lipid compositions induced by inhibition of Mgll activity (which is highly expressed in the brain) not only reduces ARA-mediated inflammation and provides neuroprotection, but also promotes differentiation of oligodendrocyte precursors (OPCs) into mature oligodendrocytes(34) which is required for remyelination. Thus, repression of Mgll promotes OPC differentiation through multiple avenues. Although a couple of Mgll inhibitors are in phase I/II clinical trials for MS treatment(35), there are no approved FDA drugs that can target Mgll.

Our previous work has shown that metformin can repress expression of Mgll by stimulating aPKC-mediated phosphorylation of histone acetyltransferase CBP at Ser436 (aPKC-CBP pathway) to promote neuronal differentiation of adult subventricular zone neural precursors (SVZ NPC)(36, 37). We recently revealed that dysregulated expression of Mgll can be a biomarker for metformin-targeted therapy to correct impaired neurogenesis and spatial memory in Alzheimer’s disease using an animal model of AD(37). In addition, our recent work(27) showed that metformin can improve CNS remyelination and social cognition using a lysolecithin-induced focal demyelination model(27). We also used OPC culture to show that metformin enhanced OPC differentiation and maturation by stimulating CBP phosphorylation at Ser436(27). Our unpublished work revealed that JZL 184, an inhibitor of Mgll activity, was able to rescue differentiation deficits of *Cbp*S436A OPCs, in which the aPKC-CBP pathway was deficient (Supplementary Figure 1). All these data suggest that metformin can act via the aPKC-CBP-mediated Mgll repression to promote remyelination and improve social cognition following demyelination. Other groups have targeted the mTOR signaling pathway(38) and have since demonstrated that metformin induces lesion reduction and elevated molecular processes that support myelin recovery via direct activation of AMPK and indirect regulation of aMPK/Nrf2/mTOR pathway in oligodendrocytes with resulting improvements in motor dysfunction(39). Metformin has also been shown to increase mitofusin-2 gene expression, which is responsible for mitochondrial fusion(40), as well as stimulate aMPK activity and improve mitochondrial function to enhance remyelination in aged rats(41).

Just as white matter demyelination in animals has been associated with social interaction deficits, the same is true in humans(42). In MS specifically, white matter pathology has been linked to impairment in social cognition and multiple research groups have identified distinct patterns of functional connectivity changes associated with social cognition(43–45). Cognitive dysfunction in MS is well documented with deficits in processing speed and memory being most prominent(46). Research differs regarding whether deficits in social cognition in MS are related to cognitive abilities in these other areas(3, 47, 48). Cognitive reserve likely plays a mediating role(49). Social cognition deficits in MS have also been associated with fatigue(3, 50), depression and anxiety(50).

To summarize, DM is a common comorbidity(4) associated with worse clinical outcomes(7), including worse cognition(14–17), in PwMS. Treatments are needed to minimize these negative outcomes and metformin may be a viable option given the cognitive benefits seen in both animal studies(27) and in people with other neurological illnesses(21–23). Mgll is a potential therapeutic target(31–33). Animal studies have demonstrated that metformin represses Mgll (37), which reduces inflammation, increases neuroprotection and promotes OPC differentiation into mature oligodendrocytes(34). Intriguingly, metformin treatment in a mouse model of MS promotes OPC regeneration and CNS remyelination, accompanied with improvements in social cognition (27). Thus, the aims of the current pilot study were threefold. First, to determine whether or not metformin represses Mgll levels in humans as has been previously demonstrated in our group’s animal work. Second, to determine if Mgll levels correlate with cognition and/or social cognition. Third, to determine if cognition differs between those with MS and comorbid diabetes treated with metformin, those with MS and comorbid diabetes not treated with metformin, those with MS and no diabetes, and healthy controls. To address these aims we enrolled a convenience sample of individuals with MS who were prescribed metformin by their treating medical team given comorbid diabetes. The overarching goal was to establish whether or not there was sufficient evidence to pursue a randomized clinical trial to evaluate the efficacy of metformin on metabolic, imaging and clinical outcomes.

## 2. Methods

For this study, we followed the STROBE case-control guidelines (see Supplementary Material for the reporting checklist)(51). Efforts were made to control for potential sources of bias. These included the inclusion of four groups while controlling for diagnoses (i.e., both DM and MS) and ensuring that all participants had an equal chance of being included (selection bias), reporting both positive and negative results (publication bias), ensuring that participants were not pre-selected for Mgll levels (observation bias), and having two independent raters checking for data entry accuracy (measurement bias).

### 2.1 Study population

A PRISMA flow diagram indicates recruitment progress (Supplementary Figure 2). Three groups of participants were recruited from the Ottawa Hospital MS Clinic, those with MS and comorbid diabetes treated clinically with Metformin (MSDM-Met), those with MS and comorbid diabetes not treated clinically with Metformin (MSDM-noMet), and those with MS and no comorbid diabetes (MSND). Healthy controls (HC) were recruited from the community, family/friends of MS participants (no first-degree relatives), and through word of mouth. Attempts were made to match the groups on age, sex, education and MS disease duration. Recruitment challenges resulted in having to control for some of these variables statistically. Sample size was restricted given that it was a convenience sample based on clinical characteristics. Inclusion criteria included age between 18 and 70 years, English-speaking, and for the MS groups, a confirmed diagnosis of MS by the 2017 McDonald criteria and EDSS between 0.0 and 8.0. Those in the MSDM-Met group must have been stably receiving Metformin treatment for at least 6 months before study participation. Those in the MSDM-noMet group were not excluded if they received Metformin treatment in the past but must have had no exposure to metformin over the preceding 6 months. These latter criteria were established based on extrapolations from animal literature that indicates changes to Mgll expression with metformin administration were observed within 56 days(37). Exclusion criteria included the presence of any neurological, medical or psychiatric condition (aside from depression or anxiety) that may have impeded cognition (e.g., prior brain injury/concussion, learning disability, attention deficit hyperactivity disorder, substance use disorder). Exclusion criteria also included an MS relapse in the preceding 30 days (for those in the MS groups), hearing or visual impairment that would have impeded their ability to see or hear test stimuli, or consumption of opioids, anxiolytics, cannabis, alcohol, caffeine or sedating medications in the 24-hours preceding the assessment.

### 2.2 Procedures

The study was approved by the Ottawa Health Science Network Research Ethics Board (20210296-01H). Participants were compensated for their parking costs but did not receive any incentives for participation. After undergoing appropriate informed consent procedures and a demographic interview, participants underwent a neuropsychological battery which included measures of cognition, emotion recognition (an aspect of social cognition), and patient-reported outcomes. The assessment took approximately 2 hours and was administered by a research assistant trained by a licensed Clinical Neuropsychologist. All groups underwent the same test battery which was administered in a fixed order to ensure standardized administration and appropriate delay intervals for memory tests. Following the assessment, a phlebotomist drew two vials of venous blood (∼10mL) from the participant and the sample was immediately delivered to a lab technician who processed the sample by gradient centrifugation. Following isolation of the blood components all samples were frozen at -80°C and later analyzed using real-time quantitative polymerase chain reaction (RT-PCR) to assess Mgll levels.

### 2.3 Measures

#### Advanced Clinical Solutions (ACS) - Test of Premorbid Functioning (TOPF)(52)

This is a measure designed to estimate premorbid intellectual functioning and can be considered a proxy measure of cognitive reserve. It requires participants to pronounce increasingly challenging phonetically irregular words. Note that Ottawa is a largely bilingual community and so both English and French pronunciations were accepted. A total raw score from 0 to 70 was recorded.

#### Brief International Cognitive Assessment for MS (BICAMS)

The BICAMS battery includes a measure of processing speed, the oral Symbol Digit Modalities Test (SDMT)(53) (3” and 2” version), as well as both verbal and non-verbal learning and memory measures respectively; the California Verbal Learning Test – II (CVLT-II)(54) and the Brief Visuospatial Memory Test – Revised (BVMT-R)(55) (published versions). Both learning and delayed recall trials for these latter two measures were administered. The BICAMS battery has been described elsewhere and thus will not be discussed in detail here(56). Canadian normative data was utilized for the BICAMS(57, 58).

#### Advanced Clinical Solutions (ACS) – Social Perception (SP)(59)

The ACS-SP assesses both basic and more complex emotion recognition from faces and voices, including higher order meaning such as sarcasm. It includes three tasks. The *Affect Naming* task requires participants to match the facial expression in a photograph of a face to an emotion from a given written list. The *Prosody-Face Matching* task requires participants to listen to a one-sentence audio clip and match the prosody depicted in the clip to one of six pictured facial expressions. The *Prosody-Pair Matching* task requires participants to match a similar audio clip to one of 6 photos of two people interacting. The participant must describe the emotion of the speaker and determine if the tone of voice changed the literal meaning of the comment (as sarcasm would). If the response is yes, the participant must then describe the true meaning of the comment. Various sub-scores can be obtained by considering these three subtests in isolation or combination. See Figure 1 for a visual depiction of how these sub-scores are derived. For the purposes of this paper, we utilized the Social Perception Total score which is derived from the total scores from each of the three sub-tests.

**Figure 1.**
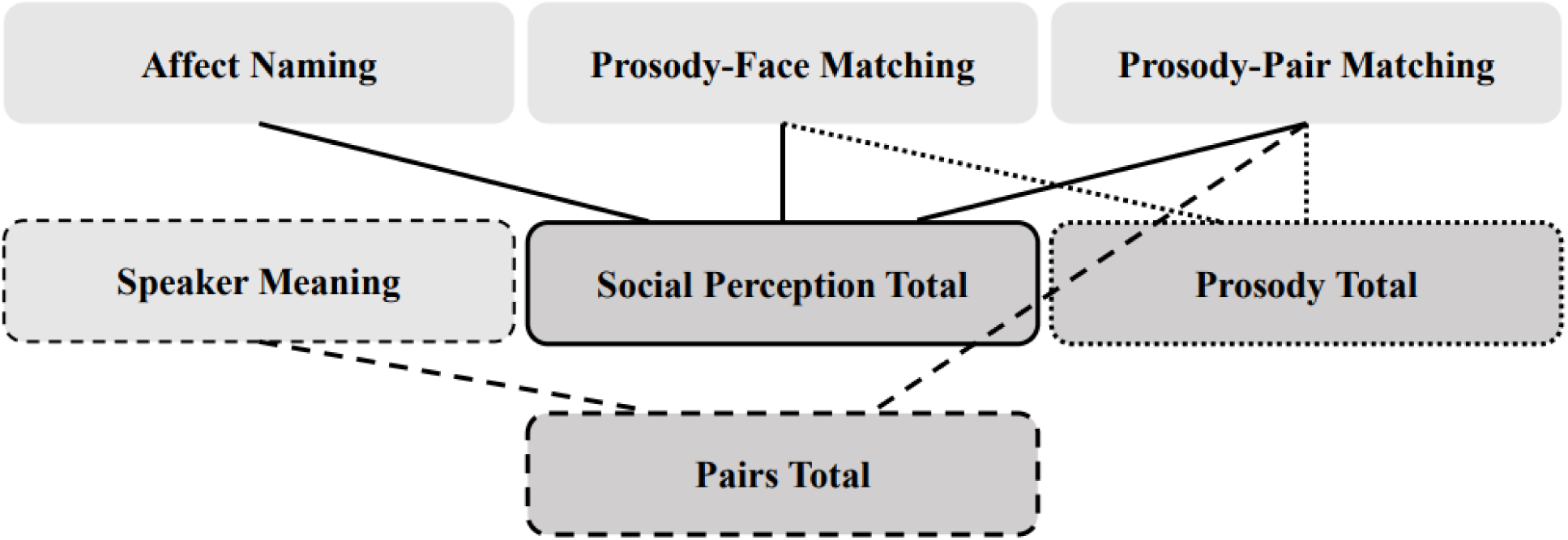
Visual Representation of the Individual and Total Scores of the Advanced Clinical Solutions Social Perception Subtests. The light boxes represent individual tasks, and the darker boxes represent total scores that are derived from these tasks. The total scores are the sum of the individual tasks to which they are connected by a line.

#### Modified Fatigue Impact Scale (mFIS)

he mFIS is a modified 21-item version of the 40-item Fatigue Impact Scale (60) that measures the effects of fatigue on physical, cognitive and psychosocial functioning. Respondents indicate how much fatigue has impacted them in the preceding month using a Likert scale (0 = no problem to 4 = extreme problem). For the current project, we utilized the total score that encompasses all three subscales. Higher scores are indicative of greater fatigue.

#### Hospital Anxiety and Depression Scale (HADS)(61)

The HADS is a 14-item self-report scale designed to detect symptoms of anxiety and depression experienced over the preceding week, including the day of assessment. There are 7 anxiety items and 7 depression items that respondents rate on a scale of 0 to 3. Higher scores correspond with greater anxiety and depression.

### 2.4 Mgll Gene Expression Analysis

Using ribonucleic acid (RNA) derived from white blood cells isolated from human blood samples, real-time quantitative polymerase chain reaction (RT-PCR)(62, 63) was used to quantify Mgll gene expression. Human blood was first separated to obtain white blood cells (WBC) using Stem Cell Technologies Lymphoprep (07851) and RNA was isolated and purified from white blood cells using the PureLink® RNA Mini Kit (12183018A). From the extracted RNA, complementary-DNA (cDNA) was synthesized using the Qiagen QuantiTect Reverse Transcription Kit (205311). Examination of Mgll expression was conducted using validated TaqMan™ Gene Expression probes targeting human Mgll, and human Glyceraldehyde 3-phosphate dehydrogenase (GADPH) as the housekeeping control. Threshold of RT-PCR expression was determined automatically by Agilent AriaMx HRM qPCR Software. Analysis of gene expression was conducted using a standard curve for quantification. Mgll expression reported is relative to the housekeeping gene GAPDH in this study.

### 2.5 Analyses

All analyses were conducted using SPSS (version 29). Analyses were conducted on available data in a pairwise fashion. Chi-square analyses determined if there were any group differences for sex or disease subtype between groups. Independent samples t-tests determined if group differences were present for other demographic variables including age, education, disease duration, EDSS and premorbid functioning. Independent samples t-tests determined if group differences were present in Mgll levels. Pearson correlational analyses determined if Mgll levels correlated with cognitive functioning. Analyses of covariance, controlling for the potential impact of premorbid ability, mood, fatigue, as well as age and education where applicable, were conducted to determine if there were group differences in cognition and emotion recognition.

## 3. Results

### 3.1 Demographic characteristics

Demographic information on the four groups can be found in Table 1. There were no group differences in terms of sex, MS subtype or EDSS. Disease duration was significantly different between the MSDM-Met and MSND groups, with the former having had MS for a greater number of years (*t* = 2.08, df = 24, *p* = .05). There were no other group differences in disease duration. Age and education were significantly different between the MSDM-Met and HC groups with the MSDM-Met group being older (*t* = 2.10, df = 36, *p* = .04) and less educated (*t* = -2.11, df = 36, *p* = .04) than the HC group. Future analyses using these groups controlled for these variables.

**Table 1.**
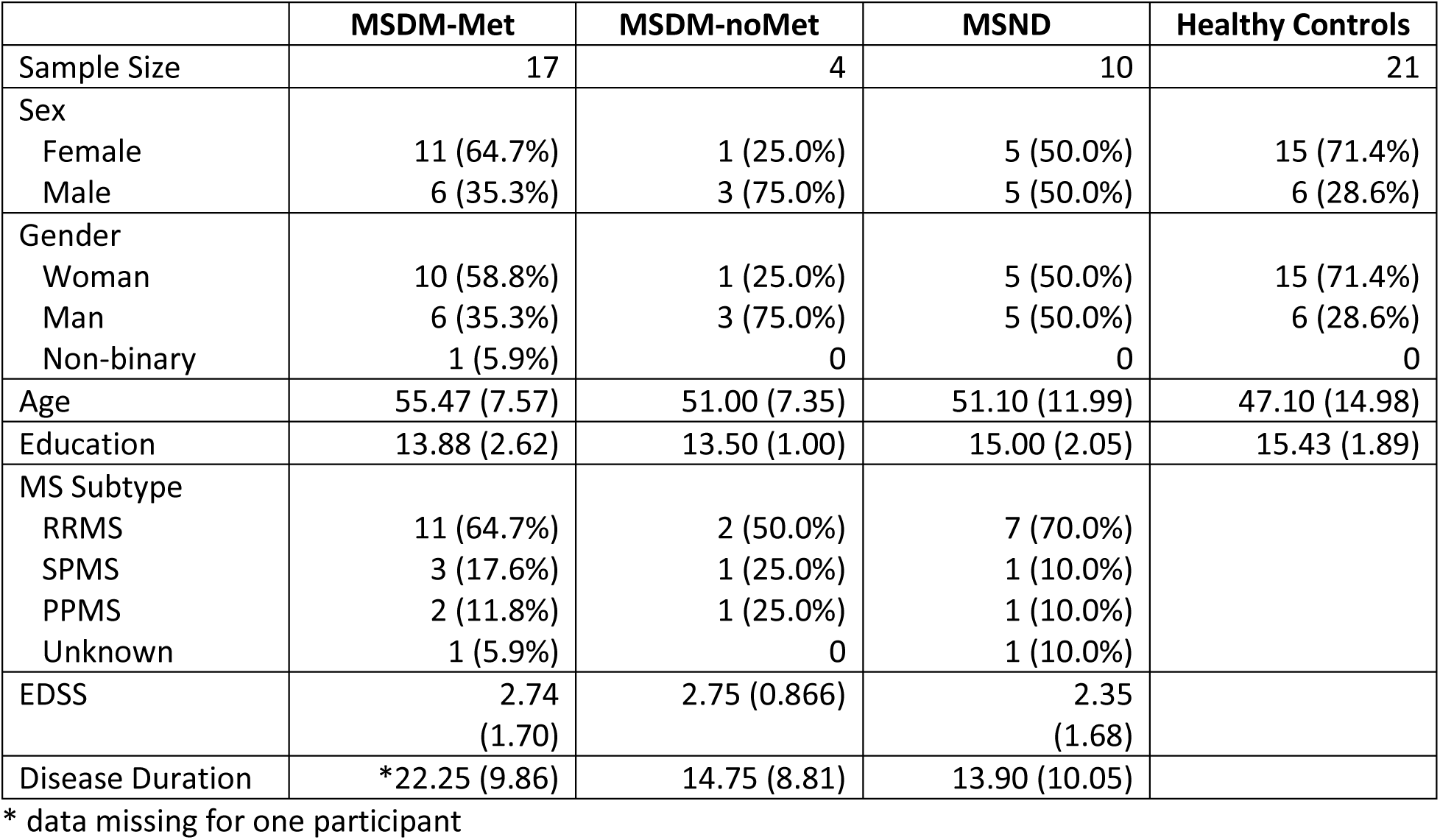
Demographic information.

|  | MSDM-Met | MSDM-noMet | MSND | Healthy Controls |
| --- | --- | --- | --- | --- |
| Sample Size | 17 | 4 | 10 | 21 |
| Sex |  |  |  |  |
| Female | 11 (64.7%) | 1 (25.0%) | 5 (50.0%) | 15 (71.4%) |
| Male | 6 (35.3%) | 3 (75.0%) | 5 (50.0%) | 6 (28.6%) |
| Gender |  |  |  |  |
| Woman | 10 (58.8%) | 1 (25.0%) | 5 (50.0%) | 15 (71.4%) |
| Man | 6 (35.3%) | 3 (75.0%) | 5 (50.0%) | 6 (28.6%) |
| Non-binary | 1 (5.9%) | 0 | 0 | 0 |
| Age | 55.47 (7.57) | 51.00 (7.35) | 51.10 (11.99) | 47.10 (14.98) |
| Education | 13.88 (2.62) | 13.50 (1.00) | 15.00 (2.05) | 15.43 (1.89) |
| MS Subtype |  |  |  |  |
| RRMS | 11 (64.7%) | 2 (50.0%) | 7 (70.0%) |  |
| SPMS | 3 (17.6%) | 1 (25.0%) | 1 (10.0%) |  |
| PPMS | 2 (11.8%) | 1 (25.0%) | 1 (10.0%) |  |
| Unknown | 1 (5.9%) | 0 | 1 (10.0%) |  |
| EDSS | 2.74<br>(1.70) | 2.75 (0.866) | 2.35<br>(1.68) |  |
| Disease Duration | *22.25 (9.86) | 14.75 (8.81) | 13.90 (10.05) |  |

\* data missing for one participant

### 3.2 Mgll levels

There were no group differences in Mgll levels. See Table 2 for the Mgll levels for each group. Nonetheless, there was a trend in the expected direction with metformin appearing to suppress Mgll levels as demonstrated in mouse models (36, 37) (See Figure 2). Given the small sample sizes, we elected to combine the MSDM-noMet group (n = 4) with the MSND group (n = 10) to determine if metformin treatment impacted Mgll levels regardless of diabetes status. When these groups were combined (i.e., all participants with MS who were not taking metformin – allMS-noMet) and compared with the MSDM-Met group (n = 17), there was a significant difference in Mgll levels in the expected direction (*t* = -2.07, df = 19, *p* = .05), suggesting that using metformin suppresses Mgll as it does in the animal literature using the measurement of human WBC gene expression. Correlational analyses revealed no significant relationship between Mgll and any of the cognitive or emotion recognition variables.

**Figure 2.**
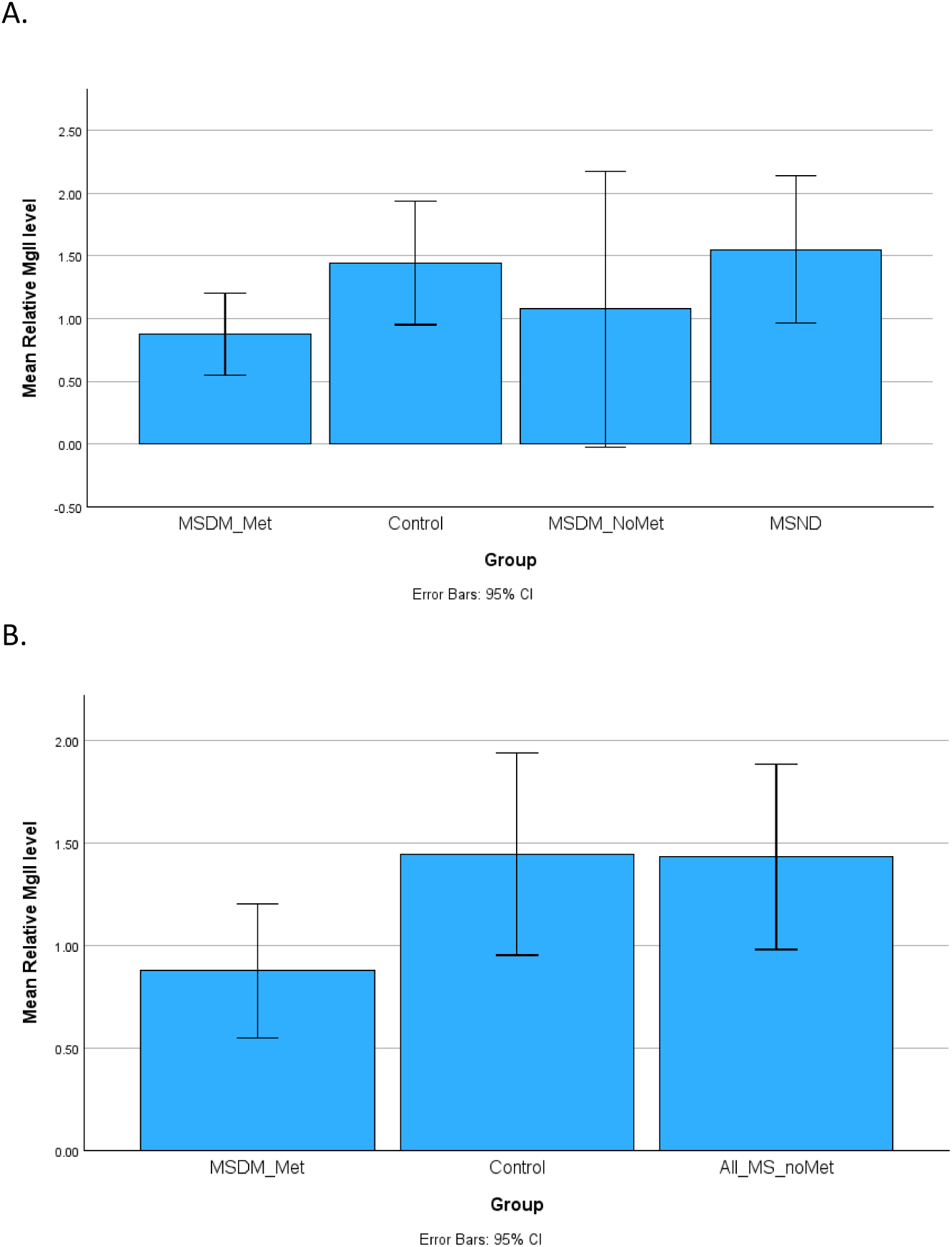
Mean Relative Mgll Levels by Group. A) Relative Mgll levels by group, B) Relative Mgll levels by group when all participants with MS not taking metformin are grouped together (MSDM-NoMet and MSND combined).

**Table 2.**
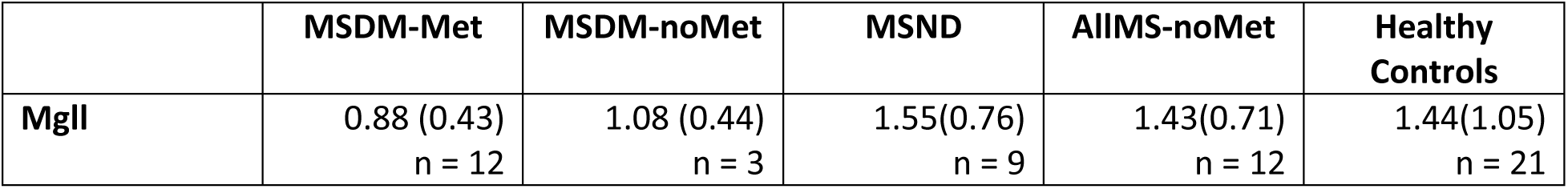
Mean Relative Mgll Levels by Group.

|  | MSDM-Met | MSDM-noMet | MSND | AllMS-noMet | Healthy Controls |
| --- | --- | --- | --- | --- | --- |
| **MgII** | 0.88 (0.43)<br>n = 12 | 1.08 (0.44)<br>n = 3 | 1.55(0.76)<br>n = 9 | 1.43(0.71)<br>n = 12 | 1.44(1.05)<br>n = 21 |

### 3.3 Cognition

Given that Mgll did not correlate with cognition, the MS groups were divided into those with diabetes (MSDM) and those without diabetes (MSND). These two groups were then compared against each other and healthy controls (see Table 3). There were no group differences when comparing MSDM to those with MSND. Similarly, there were no group differences between MSND and HC. However, when comparing the MSDM and HC groups significant differences were noted for verbal learning (*F* (1,35) = 5.85, *p* = .02, *ƞ^2^* = .14) and memory (*F* (1,35) = 5.62, *p* = .02, *ƞ^2^* = .14).

**Table 3.**
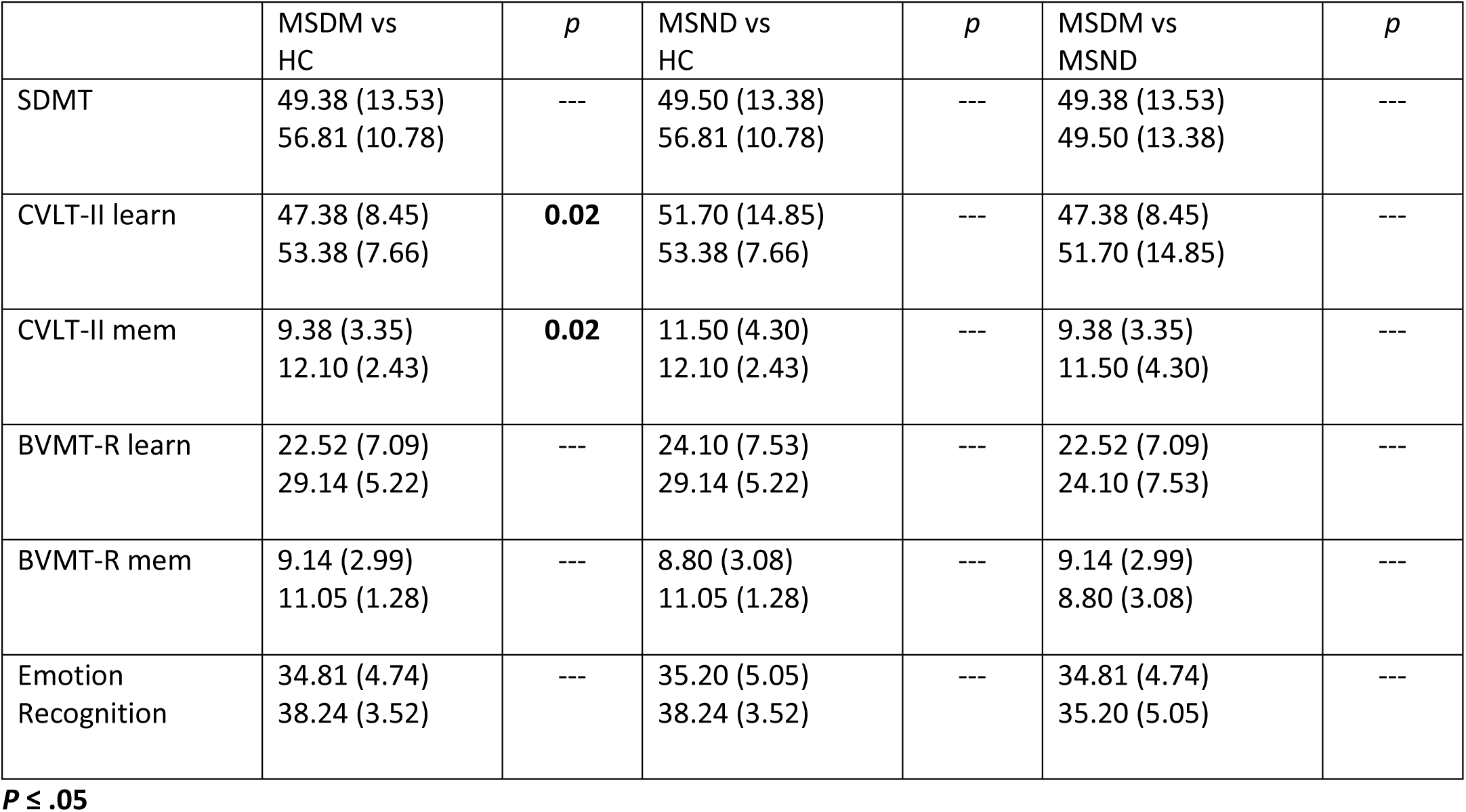
Significance values (*p*) from group comparisons for cognitive and emotion recognition measures.

| | MSDM vs HC | $p$ | MSND vs HC | $p$ | MSDM vs MSND | $p$ |
| --- | --- | --- | --- | --- | --- | --- |
| SDMT | 49.38 (13.53)<br>56.81 (10.78) | --- | 49.50 (13.38)<br>56.81 (10.78) | --- | 49.38 (13.53)<br>49.50 (13.38) | --- |
| CVLT-II learn | 47.38 (8.45)<br>53.38 (7.66) | **0.02** | 51.70 (14.85)<br>53.38 (7.66) | --- | 47.38 (8.45)<br>51.70 (14.85) | --- |
| CVLT-II mem | 9.38 (3.35)<br>12.10 (2.43) | **0.02** | 11.50 (4.30)<br>12.10 (2.43) | --- | 9.38 (3.35)<br>11.50 (4.30) | --- |
| BVMT-R learn | 22.52 (7.09)<br>29.14 (5.22) | --- | 24.10 (7.53)<br>29.14 (5.22) | --- | 22.52 (7.09)<br>24.10 (7.53) | --- |
| BVMT-R mem | 9.14 (2.99)<br>11.05 (1.28) | --- | 8.80 (3.08)<br>11.05 (1.28) | --- | 9.14 (2.99)<br>8.80 (3.08) | --- |
| Emotion Recognition | 34.81 (4.74)<br>38.24 (3.52) | --- | 35.20 (5.05)<br>38.24 (3.52) | --- | 34.81 (4.74)<br>35.20 (5.05) | --- |

$P \leq .05$

## 4. Discussion

### 4.1 Monoacylglycerol Lipase (Mgll)

The first aim of this study was to determine if metformin represses Mgll levels in humans as was previously demonstrated in our group’s animal work(27). When comparing participants with MS who were not taking metformin to those who were taking metformin, the latter group had lower Mgll levels as expected. This suggests that metformin indeed suppresses Mgll as it does in the animal literature(36, 37) and provides preliminary support for metformin as a potential therapeutic agent in MS. Despite the expected and promising response in the underlying biological processes, Mgll did not correlate with cognition or emotion recognition in humans. This is in contrast to animal models that found clinically relevant implications of metformin treatment, potentially causing Mgll suppression(37), in the form of improved social interaction in a mouse model of MS (27), and of Mgll suppression and improved cognition in a mouse model of chronic traumatic encephalopathy(64). A recent review of studies evaluating Mgll suppression in Alzheimer animal models noted inconsistent findings(65), suggesting that much remains unknown about the potential cognitive implications of Mgll suppression. The following questions remain unaddressed: 1. How much Mgll suppression is required to produce clinical benefits? 2. Does metformin dose matter? 3. What is a typical Mgll level in the general population? 4. What naturally occurring factors affect Mgll levels? While the present study suggests that Mgll levels are comparable between PwMS and healthy controls (see Figure 2B), there was a high level of individual variability; thus, it remains unclear which factors might have influenced these levels.

Methodological differences may also explain why Mgll did not correlate with cognition in the current sample. The present sample size was smaller than initially anticipated (see below) and this may have been a factor that resulted in a lack of any observable relationship. Alternatively, the lack of a relationship may have been related to how cognition was assessed. In our group’s animal model, social cognition was evaluated by examining the duration of time a mouse spent in close proximity to another mouse (i.e., amount of social interaction)(27). Given that these same parameters are not easily replicated in a human clinic setting, the present study opted to evaluate social cognition more traditionally by using a task of emotion recognition, a method previously established in the human MS literature(66). The lack of equivalency between these two methods with respect to how they conceptualize social cognition may have accounted for the dissimilar outcomes. It is conceivable that Mgll suppression in humans could have impacted the degree of social interaction or a person’s desire to remain socially engaged, despite any clear impact on their emotion recognition. Future studies, therefore, should consider examining social network size or degree of socialization in addition to emotion recognition. Initial work has demonstrated that social network size in PwMS is related to other aspects of cognition (i.e., processing speed and memory)(67, 68) and is an emerging area of interest.

### 4.2 Cognition and Emotion Recognition

When comparing those with MS and DM, those with MS alone, and healthy controls, group differences in cognition were only found between the MSDM group and healthy controls. Specifically, the MSDM group demonstrated poorer verbal learning and memory as measured by the CVLT-II total learning score and the long-delay free recall score. There were no other group differences found. This finding of lower verbal learning and memory in those with MS and comorbid DM is consistent with a previous study demonstrating that a higher number of vascular comorbidities is associated with lower cognitive function, including lower scores on the CVLT-II total learning score(16). The cognitive differences were explained by differences in brain structure, suggesting that vascular comorbidity may influence cognitive function via increased peripheral inflammation, endothelial injury, and alterations in blood vessel function, cerebral blood flow and metabolism(16).

That the present study did not find a negative impact on cognition for those with MS alone is in contrast with well-established findings in the research literature that demonstrate poorer cognition in those with MS compared to healthy controls(13). The present study did not have a DM-only group, but a similar negative impact on cognition would have been expected as a result of DM alone given the increased risk of cognitive impairment with diabetes(69). The lack of significant cognitive differences resulting from MS alone in the current study is likely reflective of our small sample size, and the subsequent lack of statistical power. Despite this, examination of the mean scores on all cognitive and facial recognition measures (see Table 3) revealed that those with MS did tend to perform uniformly poorer than healthy controls, indicating a trend in the expected direction, though not to a statistically significant degree. The small sample size also likely accounted for the lack of group differences in emotion recognition given that the mean values were also indicative of a trend for lower performance in the MSDM and MSND groups compared to controls. Recent systematic reviews have demonstrated a negative impact of MS on social cognition and emotion recognition more specifically(70, 71), and examination of social cognitive variables is gaining more attention in the literature. Our current findings suggest that neither diabetes nor MS alone are driving the differences in verbal learning and memory, but rather, it is the combination of the two illnesses together that lead to cognitive impairment of sufficient magnitude to be detected even in this small sample.

### 4.3 Limitations

The present study was not designed as a treatment trial. The goal was to evaluate whether metformin impacted Mgll levels and cognition (including social cognition) in the same manner as was demonstrated in animal models(27) to determine if a treatment trial is justified. Thus, the decision to enroll participants with diabetes was made given that this is a convenience population likely to be treated clinically with metformin. This method of recruitment resulted in two limitations. First, given that the incidence of diabetes in the MS population is comparable to the general population (72), we anticipated that recruitment of 30 participants per MSDM group (i.e., 30 on metformin and 30 treated with diet or other diabetes medications) would be quite feasible. Unfortunately, in addition to recruitment being negatively impacted by COVID pandemic-related delays, we soon realized that metformin is the primary first line treatment for diabetes despite newer classes of medications being available(73), and thus the pool of individuals available who were not taking metformin was very small. This resulted in sample sizes being lower than planned. Second, enrollment of individuals with both MS and comorbid diabetes means that we cannot generalize the current findings to the MS population as a whole. We attempted to address this by enrolling an MS-only group, but sample sizes remained small and thus conclusions were tempered accordingly.

## 5. Conclusions

As was anticipated, metformin suppresses Mgll in humans as previously documented in animal literature. This lends additional support for further investigation of metformin as a potential treatment for MS given the demonstrated impact in animal models on cellular processes via decreased inflammation, increased neurogenesis and enhanced oligodendrocyte precursor differentiation. However, Mgll does not correlate with cognition in humans possibly due to sample size or methodological differences between animal and human studies. Future studies should evaluate the potential impact of metformin treatment on social network size or desire to remain socially engaged to align more closely with outcomes in animal studies. Questions remain regarding Mgll in humans, such as what are normal levels of Mgll, and what degree of change is required to lead to biological and clinical change? Current results also reveal that the combination of MS and DM together has negative implications for both verbal learning and memory, supporting emerging literature demonstrating that vascular comorbidity increases the risk of cognitive dysfunction in people with MS. Findings support the pursuit of a larger clinical trial evaluating efficacy of metformin for biological, imaging and clinical outcomes. Finally, the current findings support the monitoring of comorbidities in those with MS. Clinicians should strongly advocate for healthy lifestyles (e.g., exercise, improved nutrition) to prevent DM and other vascular comorbidities.

### Statements and Declarations

The study was approved by the Ottawa Health Science Network Research Ethics Board (20210296-01H).

## Data Availability

All data produced in the present study are available upon reasonable request to the authors.

## Acknowledgements

The authors would like to thank the people living with MS and the healthy control participants who participated in this study. Their time and effort are much appreciated.

## Availability of Data and Material

If researchers would like to obtain data, they can contact the first author directly at.

