## Supplemental Figure 1 for "Metformin, monoacylglycerol lipase expression, cognition and emotion recognition in people with multiple sclerosis and comorbid type II diabetes: A case-control study"

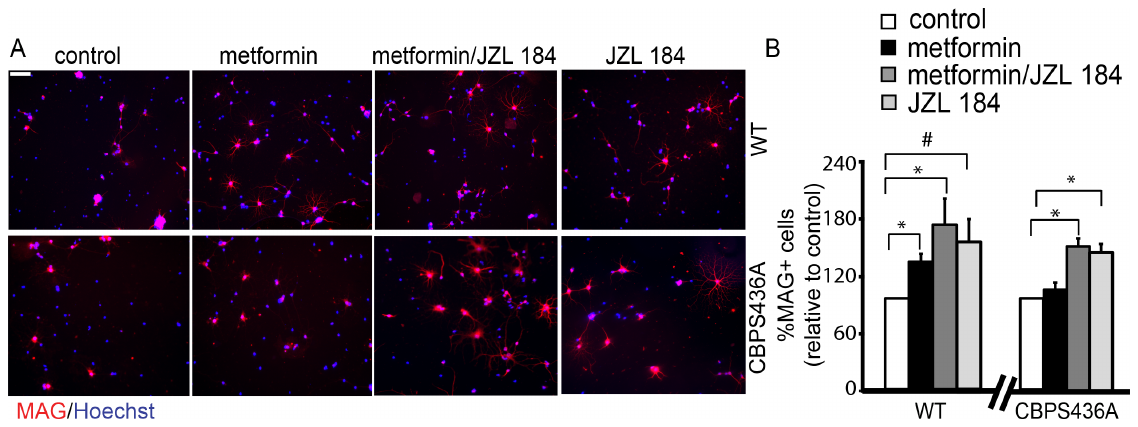

**Suppl Figure 1:** **Metformin promotes OPC differentiation through activating the aPKC-CBP mediated Mgll repression.** (A-B) Photographs and quantitative analysis of the percentage of MAG+ immature oligodendrocytes from the enriched OPCs isolated from postnatal 0-2 days (P0-2) )wild type and CbpS436A pups, cultured in the absence and presence of metformin (10 µM) and treated with vehicle (control) or JZL 184 (1 µM), and immunostained for MAG (red) and Hoechst (blue). Scale bar: 80 µm. (n=4/group, *p < 0.05, #p=0.05).
