## Supplemental Figure 2 for "Metformin, monoacylglycerol lipase expression, cognition and emotion recognition in people with multiple sclerosis and comorbid type II diabetes: A case-control study"

*Supplementary Figure 2: PRISMA Flow Diagram For Each Stage of Study*

Identification

MS clinic records identified through automatized Epic database searching

MS: *n* = 233

Additional records identified through manual searching & word-of-mouth

MS: *n* = 32

Healthy Controls: *n* = 21

Screening

Records excluded

MS: *n* = 227

Records screened for eligibility criteria

MS: *n* = 265

Healthy Controls: *n* = 21

Enrollment

Records excluded

MS: *n* = 5

5 scheduling conflicts

Number of Participants Enrolled

MS: *n* = 38

Healthy Controls: *n* = 21

Consent

Number of Participants Consented

MS: *n* = 33

Healthy Controls: *n* = 21

Final Number of Participants Included

MS: *n* = 31

Healthy Controls: *n* = 21

Records excluded

(n = 2)

1 scheduling conflict

1 Informed study team of a learning disability following study participation

Included

Number of Participants Included in Analyses

MS: *n* = 33

Healthy Controls: *n* = 21
